# Defining the rhythm: a new method to classify tremor and myoclonus

**DOI:** 10.1101/2025.04.13.25325747

**Authors:** Anna Latorre, Blake Hale, Carla Cordivari, Kais Humaidan, John C. Rothwell, Kailash P. Bhatia, Lorenzo Rocchi

**Affiliations:** Department of Clinical and Movement Neurosciences, UCL Queen Square Institute of Neurology, University College London, London, United Kingdom; Department of Clinical Neurophysiology, National Hospital for Neurology and Neurosurgery, University College London Hospitals, Queen Square, London, United Kingdom; Department of Medical Sciences and Public Health, University of Cagliari, Cagliari, Italy

**Keywords:** essential tremor, orthostatic tremor, dystonic tremor, myoclonus, power spectral density analysis

## Abstract

**Background:** Tremor hallmark feature is rhythmicity, which can be quantified using power spectral density analysis. However, tremor exhibits considerable variability, ranging from highly regular to more irregular patterns. Similarly, rhythmicity in myoclonus also varies, but it typically manifesting as arrhythmic jerks.

**Objectives:** To develop power spectral density-based measures of movement regularity for the classification tremor and myoclonus.

**Methods:** Electromyography data from 153 patients were analysed retrospectively, including orthostatic tremor (n=36), essential tremor (n=40), dystonic tremor (n=42), and limb cortical myoclonus (n=35). Five power spectral density analysis-derived variables were assessed: peak prominence, peak-to-broadband power ratio, peak frequency, peak width, and harmonics. Discriminant analysis evaluated classification accuracy across groups.

**Results:** Peak prominence was highest in orthostatic tremor and higher in essential tremor than dystonic tremor or myoclonus. Peak-to-broadband power ratio showed similar trends. Peak frequency differed across groups, with myoclonus highest and orthostatic tremor exceeding essential tremor and dystonic tremor. Peak width was larger in myoclonus and, to a less extent, in dystonic tremor compared to essential tremor. Harmonics were greater in orthostatic tremor and essential tremor compared to dystonic tremor and myoclonus. Discriminant analysis correctly classified 86.3% of cases, with overlap between essential tremor and dystonic tremor. Receiver operating characteristic curve analysis for peak prominence and width demonstrated high classification accuracy between essential tremor and dystonic tremor.

**Conclusions:** Our findings represent a promising initial step toward establishing objective, power spectral density-based measures for the classification of tremor and myoclonus. These tools could enhance diagnostic accuracy and deepen insights into these disorders.

## Introduction

Tremor is defined as an involuntary, rhythmic, oscillatory movement of a body part ^1^. Its hallmark feature, rhythmicity, distinguishes it from other hyperkinetic movement disorders, providing a valuable diagnostic advantage. This regular muscle discharges can be readily identified using basic electrophysiological techniques such as surface electromyography (EMG). However, tremor rhythmicity may range from highly regular to somewhat irregular ^2–4^. While regularity is a key feature of tremor, it can also vary in myoclonus. Certain forms of cortical and high-frequency myoclonus may exhibit rhythmicity that visually resembles tremor, complicating the clinical distinction between the two conditions ^5^. Aside from these exceptions, myoclonus is typically considered the opposite of tremor, as it is usually characterized by arrhythmic jerks ^5, 6^.

Despite advances in understanding tremor and myoclonus, the translation of neurophysiological concepts into clinical practice remains limited. A key challenge lies in identifying neurophysiological characteristics that accurately reflect observable rhythmicity. The absence of standardized, objective metrics has led to imprecise clinical terminology. For instance, terms such as “jerky” are often used to describe “irregular” tremor - a description that may appear contradictory. This term is frequently applied to dystonic tremor (DT), yet it introduces ambiguity, as “jerk” is synonymous with myoclonus. Describing tremor as “jerky” could imply the presence of brief, abrupt muscle contractions (i.e., myoclonus) superimposed on the tremor, further complicating differentiation between these phenomena.

A well-established method for quantifying movement rhythmicity, as recorded via EMG or accelerometery, is the estimation of power spectral density (PSD) using the Fast Fourier Transform or related techniques. The presence of a peak in the PSD is often regarded as a reliable indicator of rhythmicity; however, no objective definition of a PSD peak currently exists ^4^. When visually apparent, peak frequency can aid in diagnosing tremors with distinct frequencies, such as orthostatic tremor (OT) and essential tremor (ET) ^1, 7^. Conversely, for DT or myoclonus, the presence of a PSD peak is less well-studied and may be ambiguous or absent.

Beyond identifying peaks, more sophisticated PSD-derived measures remain underexplored ^8, 9^. One such feature, peak width, reflects the range of frequencies contributing to the tremor signal and it has been proposed to distinguish ET from ET-plus and DT ^10, 11^. A broader peak width may correspond to greater irregularity observed during visual inspection, though this relationship has yet to be systematically investigated. Other potential PSD features, such as the prominence of peak activity relative to the underlying broadband PSD, could offer valuable insights. In theory, more regular movements would exhibit less contamination from non-rhythmic activity, resulting in a more prominent peak. Additionally, a regular movement pattern may be inferred from the presence of harmonics at integer multiples of the fundamental tremor frequency. Strong synchronization of motor unit spike trains would enhance tremor regularity, resulting in pronounced harmonics in both the neural drive and the EMG signal ^12^.

To test these concepts and develop objective measures of movement regularity, we analysed four distinct conditions: OT, characterized by extreme regularity and a clear PSD peak; myoclonus, typically irregular; ET, noted for its regularity; and DT, considered less regular than ET and often described as “jerky,” though still classified as tremor. We extracted PSD-derived measures across these conditions and demonstrated that they form a continuum of regularity. Furthermore, these measures, when combined, effectively differentiate between the conditions.

## Materials and methods

### Patients

Data from 153 patients who underwent tremor or myoclonus EMG recording, at UCL Queen Square Institute of Neurology, between 2019 and 2023 were retrospectively collected and analysed. Medical records for all participants were systematically reviewed to confirm the final diagnosis. The cohort comprised 36 patients with OT, 40 with ET, 42 with DT, and 35 with cortical myoclonus, diagnosed according to the current criteria ^1, 13^. The study protocol was approved by the local institutional review board and conducted in accordance with the Declaration of Helsinki.

### EMG recording and analysis

EMG recording is detailed in the Supplementary Materials. Examples of raw EMG traces are provided in Figure 1, Panel A.

**Figure 1.**
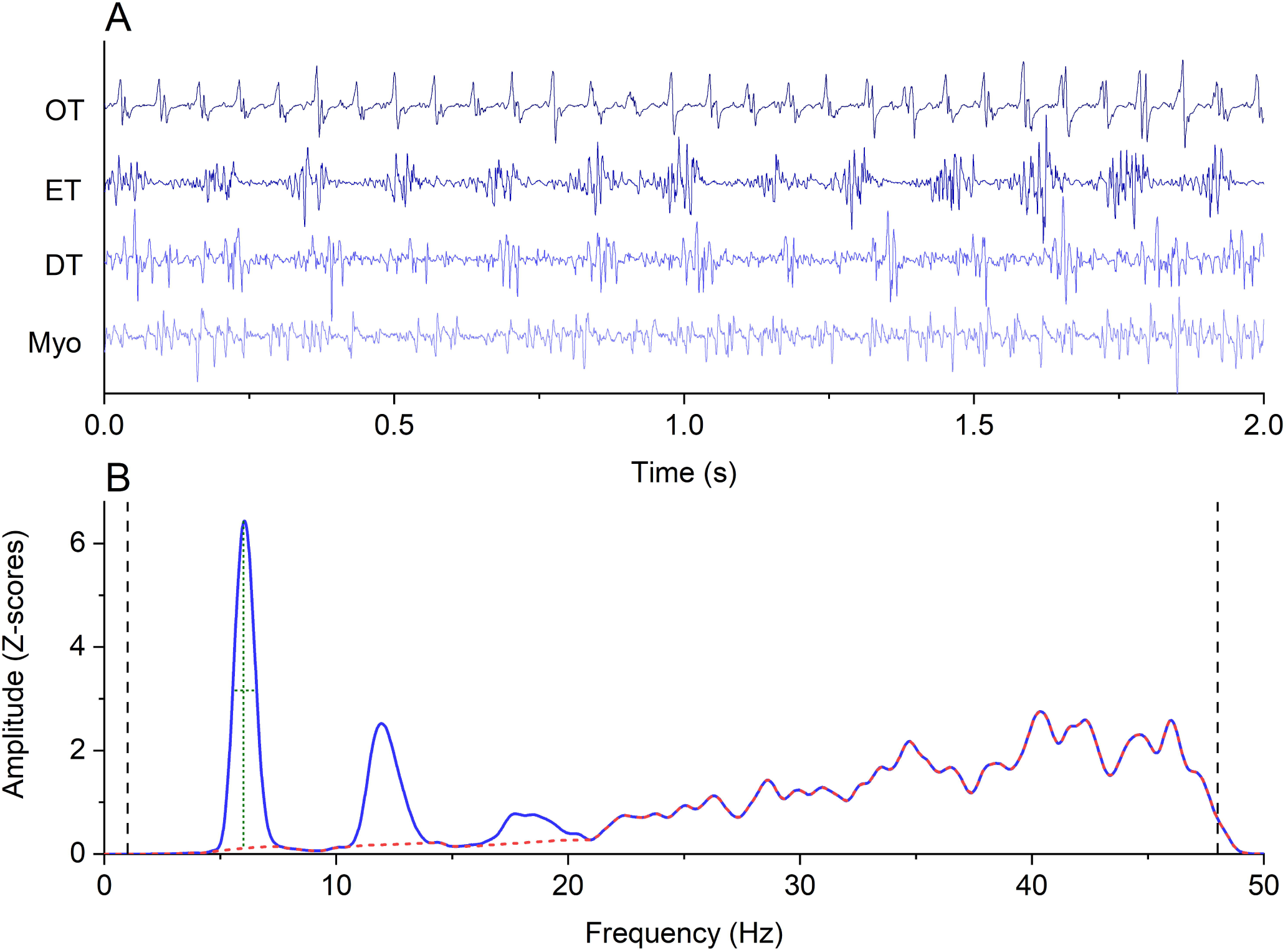
Panel A: EMG traces of the four conditions explored, showing maximum rhythmicity in OT (upper row), progressively decreasing in ET, DT and Myoclonus (Myo). Panel B: example of a PSD from a patient with ET and graphical representation of features used to extract the main variables. Black vertical dashed lines represent the range (1-48 Hz) from which broadband EMG activity, used to calculate the PB ratio, was selected. The dashed red line indicates the PSD where activity from harmonics and peak frequency was interpolated, to measure broadband activity and peak prominence (vertical dotted green line). The horizontal dotted green line represents peak width. See text for further information.

Signal analysis was performed in MATLAB (Version 2020a, MathWorks Inc., Natick, USA). For all EMG recordings, PSD was calculated using Welch’s periodogram with a Hann window twice the length of the sampling rate and a frequency resolution of 0.1 Hz. From the PSD, a peak of activity was identified by selecting the maximum value within predefined frequency ranges for each condition: 1-20 Hz for OT, 1-12 Hz for ET and DT, and 1-48 Hz for myoclonus, excluding harmonics (see below). These ranges were chosen based on the expected peak frequencies for each group while avoiding high-amplitude, non-tremor-related EMG activity from being erroneously selected. The peak detection process used MATLAB’s ‘findpeaks’ function and was visually confirmed.

Once a peak was identified, five key variables were extracted. The first variable was peak prominence, calculated as the ratio of peak power to the underlying EMG power at the same frequency. The underlying EMG power was estimated by interpolating the PSD after excluding the peak and surrounding values using a cubic function, then selecting the PSD value at the peak frequency from this interpolated spectrum. The second variable was the peak-to-broadband power ratio (PB ratio), where broadband power was defined as the average PSD between 1 and 48 Hz (excluding the peak and harmonics) using the same interpolation approach. The third variable was the peak frequency itself, while the fourth was the peak width at half prominence ^10, 11^. Finally, the fifth variable was the number of harmonics, defined as regions of the PSD at frequencies that were integer multiples of the peak frequency, where power exceeded the underlying EMG power by at least 3 standard deviations (SD), as determined using the interpolation method. A graphical explanation of these variables is provided in Figure 1, Panel B.

### Statistical analysis

Statistical analyses were carried out with SPSS version 26.0 (IBM Corp, Armonk, USA). Equality of covariance matrices was tested by means of Box’s test. A multivariate analysis of variance (MANOVA) was performed to investigate simultaneous differences across the four different patient groups (OT, ET, DT, myoclonus), which were included as a fixed factor. Dependent variables, each having four levels corresponding to the four patient groups, included peak prominence, PB ratio, peak frequency, peak width and number of harmonics.

To investigate differences in individual variables between groups, the MANOVA was followed by five univariate between-group analyses of variance (ANOVAs). For variables showing statistically significant effects, Bonferroni-corrected post hoc comparisons were conducted to identify specific group differences. Post hoc tests included t-tests for peak prominence, PB ratio, peak frequency, and peak width, while the Mann-Whitney U test was applied for the number of harmonics.

Finally, a discriminant analysis was performed to assess whether a linear combination of the five outcome variables could effectively discriminate among the four patient groups. Prior probabilities for group classification were based on the observed group sizes.

## Results

A graphical summary of the PSDs and their peak features is provided in Figure 2. Box’s test for equality of covariance matrices was not significant (p = 0.18), and Levene’s test for equality of error variances showed no significant results for any of the tested variables (all p > 0.05).

**Figure 2.**
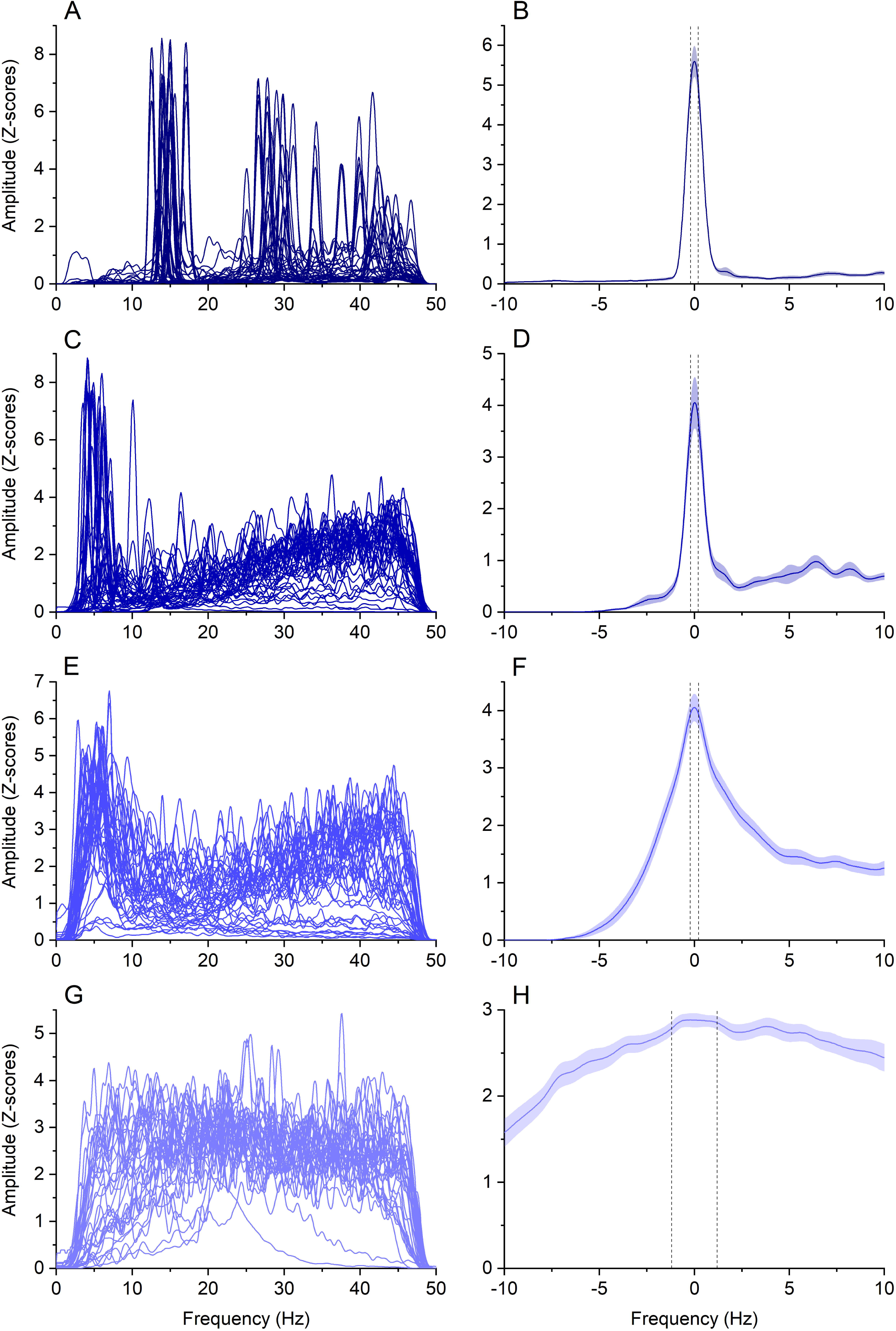
In panels A, C, E and G PSDs from all subjects from OT, ET, DT and myoclonus patients are shown, respectively. To facilitate visual comparison, voltage values were converted into Z-scores and offsets were subtracted. Panels B, D, F and H show frequency-centred peaks of activity for OT, ET, DT and myoclonus, respectively. OT exhibited narrower and more pronounced peaks in the PSD compared to other conditions. In contrast, the PSD peaks for ET, DT, and myoclonus became progressively broader and less prominent relative to surrounding EMG activity. Black dashed lines represent the standard error of the mean for frequency and shaded areas represent the standard error of the mean for the signal.

Using Pillai’s trace, the MANOVA showed a significant effect of “group” (F_15,441_ = 35.864, p < 0.001). The one-way ANOVAs all showed significant main effects of factor “group” (peak prominence: F_3,149_ = 59.351, p < 0.001; PB Ratio: F_3,149_ = 31.115, p < 0.001; Peak frequency: F_3,149_ = 141.768, p < 0.001; Peak width: F_3,149_ = 52.556, p < 0.001; Harmonics: F_3,149_ = 77.810, p < 0.001). Post-hoc comparisons on peak prominence showed significantly larger values in OT, compared to the other groups (all p values < 0.001). This was also the case for ET compared to DT and myoclonus (p = 0.001 and p < 0.001) while the difference between DT and myoclonus was not significant (p = 0.686), albeit the first showed larger values (Figure 3, panel A). Post-hoc tests on PB ratio showed similar but weaker results, with OT having significantly larger values than ET, DT and myoclonus (all p values < 0.001), while the same variable was not significantly different in the last three groups (Figure 3, panel B). The four groups showed marked differences in frequency of peak activity, with myoclonus being significantly larger than the other three (all p < 0.001) and OT showing larger values than ET and DT (both p values < 0.001). Conversely, the same variable was not significantly different when comparing ET and DT (p = 0.951) (Figure 3, panel C). Peak width was significantly larger in myoclonus compared to OT, ET and DT (all p values < 0.001) and significantly larger, albeit to a less extent, in DT compared to ET (p = 0.023) and OT (p = 0.019). The same variable, however, did not yield statistically different values comparing OT and ET (p = 0.945) (Figure 3, panel D). Post hoc comparisons on the number of harmonics showed a significantly larger value in OT, compared to the other three groups (all p values < 0.001). The same was true when comparing ET with DT and myoclonus (both p values < 0.001) while there was no significant difference between the last two groups (p > 0.05) (Figure 4).

**Figure 3.**
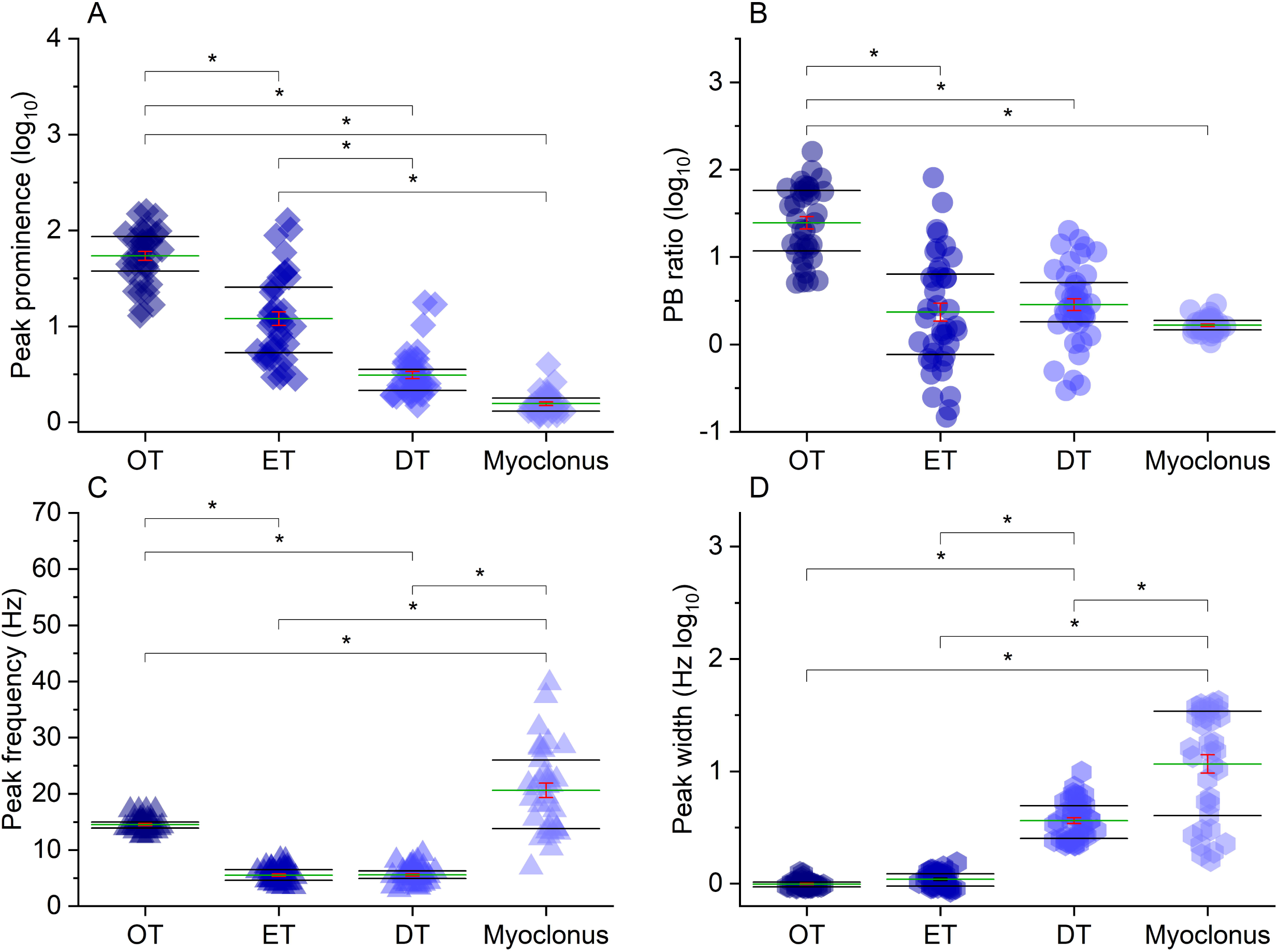
Panel A, B, C and D show post-hoc comparisons for peak prominence, PB ratio, peak frequency and peak width, respectively, for the four patient groups investigated (OT, ET, DT, myoclonus). Brackets with asterisks indicate statistically significant comparisons. To facilitate visual comparison across the four groups, Y axis values have been converted into Log_10_ values for peak prominence, PB ratio and peak width. Raw values are as follows. Peak prominence (mean ± standard deviation): OT 65.17 ± 6.24, ET 21.17 ± 4.42, DT 3.77 ± 0.53, myoclonus 1.62 ± 0.09; PB ratio: OT 37.34 ± 5.58, ET 7.1 ± 2.26, DT 4.49 ± 0.70, myoclonus 1.69 ± 0.06; Peak frequency: OT 14.53 ± 0.21, ET 5.51 ± 0.20, DT 5.56 ± 0.22, myoclonus 20.62 ± 1.28; Peak width: OT 0.99 ± 0.01, ET 1.10 ± 0.02, DT 3.94 ± 0.26, myoclonus 18.45 ± 2.41.

**Figure 4.**
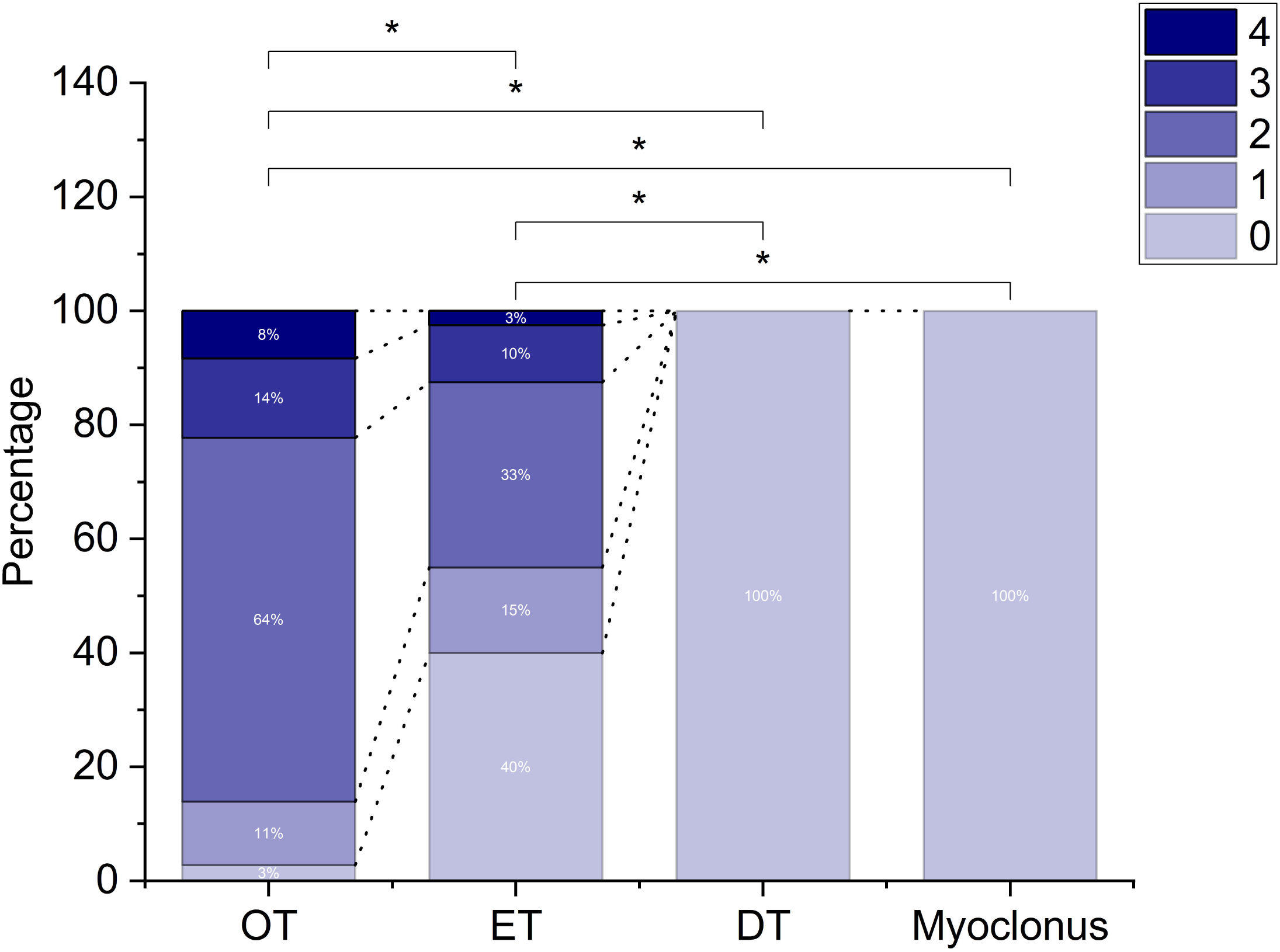
Percentages of cases in each group (OT, ET, DT, myoclonus) where variable numbers of harmonics (ranging from 9 to 4) were present. Brackets with asterisks indicate statistically significant comparisons. Black dashed lines join areas in the stacked bars corresponding to the same number of harmonics. Overall, OT showed the largest number of harmonics, followed by ET. No harmonics were observed in DT and myoclonus recording.

The discriminant analysis revealed three discriminant functions (DF), each significantly separating the four patient groups (p values < 0.001, < 0.001 and 0.002 respectively). Variance explained and canonical R^2^ were 63.3%/0.91, 35.3%/0.853 and 1.4%/0.306 for DF1, DF2 and DF3, respectively. In combination, these functions significantly differentiated the four groups (A = 0.043, χ2(15) = 465.343, p < 0.001). The correlations between outcomes and the DFs revealed that frequency and width significantly loaded on DF1 (r = 0.719 and 0.443, respectively). By contrast, peak prominence and harmonics loaded significantly on DF2 (r = 0.645 and r = 0.702, respectively), while PB ratio significantly loaded on DF3 (r = −0.674). Details about single-subject scores are depicted in Figure 5. When classified according to the DFs, 86.3% of original cases were correctly identified, specifically 97.2% of OTs (35/36, 1 identified as ET), 60% ETs (24/40, 16 identified as DTs), 100% of DTs and 97.1% myoclonus (34/35, 1 identified as DTs). The difficult discrimination between ET and DT might be due to some overlapping features, such as peak frequency and PB ratio, which did not show statistically significant differences (Figure 3) and may have decreased the accuracy of the discriminant analysis. Therefore, we further explored the possibility to discriminate ET and DT by focussing on spectral features showing statistically significant differences between these two groups (peak prominence and peak width). To this aim, we calculated receiver operating characteristic (ROC) curves separately for both variables. This analysis, shown in the Supplementary Figure, yielded high classification accuracy for both variables (area under the curve 0.905 and 0.963 for peak prominence and peak width, respectively).

**Figure 5.**
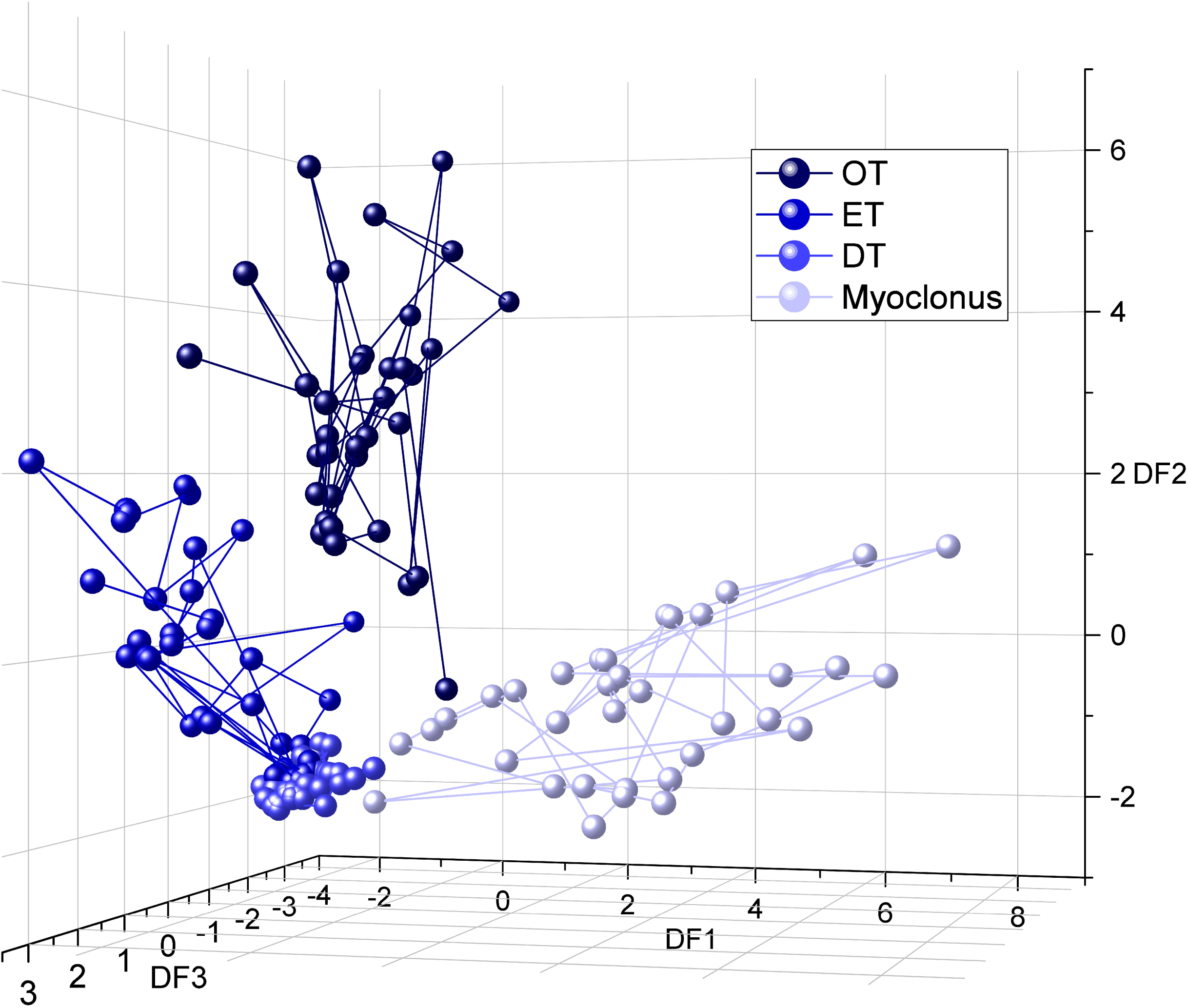
Clustering of PSD features obtained in OT, ET, DT and myoclonus according to the three discriminant functions (DF1, DF2, DF3), represented by three different axes. Overall, DF1 and DF2, which explained the largest variance, could easily discriminate OT and myoclonus from other groups, while ET and DT were the groups most difficult to separate. See text for details.

### Data availability

The data that support the findings of this study are available from the corresponding author, upon reasonable request.

## Discussion

In this study, we analysed five measures extracted from the PSD: peak prominence, PB ratio, frequency of peak activity, peak width, and number of harmonics. While the individual measures demonstrated varying results, they collectively highlighted significant differences among the tremor subtypes (OT, ET, DT) and between these and cortical myoclonus, aligning with their distinct phenomenological characteristics. Furthermore, discriminant analysis combining all outcome measures successfully separated the four patient groups, with the exception of ET and DT. However, these two groups were more effectively distinguished using peak prominence and peak width, underscoring the utility of specific spectral features for finer diagnostic differentiation.

### Peak prominence and PB ratio

The peak prominence of the PSD reflects how distinctly the most prominent signal (the peak) stands out from the background activity at a specific frequency. The PB ratio is a similar concept, but it compares the peak power to a broader measure of background activity, encompassing the average power across the entire frequency range considered (1–48 Hz). This allows the peak power to be assessed relative to the overall EMG activity, making it possible to distinguish tremor-related muscle activity (the peak) from background noise due to non-specific muscle activity. The group with the highest prominence was OT, which showed a statistically significant difference compared to the others. Conversely, the ET peak was significantly more prominent than both DT and myoclonus, while DT and myoclonus did not differ significantly, though DT displayed a larger value. The PB ratio demonstrated a similar pattern of results, albeit weaker, since only OT showed statistically significant differences with the three other groups (Figure 3, panels A and B).

These measures are relatively new, as only a few studies have applied similar approaches in this context ^8^. High synchronization among motor units at the tremor frequency has already been demonstrated in ET ^14^. However, increased peak power is also observed in the “voluntary drive frequency band,” attributed to voluntary contraction during posture holding. Additionally, peripheral feedback loops can influence motor unit discharge during tremor ^15^. This indicates that background activity at frequencies matching or differing from the tremor frequency may be related to voluntary movements, afferent inputs, or other EMG activity unrelated to tremor.

Our findings suggest that tremor in ET and OT is driven by highly synchronized motor unit discharges, as previously indicated ^7, 14, 15^. This synchronization results in a tremor-related peak in the PSD that is much larger than the EMG activity associated with posture. In contrast, in DT, dystonic activity likely reduces the gap between the peak and background noise. This suggests that only part of the dystonic activity follows a regular pattern contributing to tremor, while irregular and continuous dystonic activity persists in the background. Lastly, the findings in myoclonus align with expectations: a clear peak is absent, with irregular, chaotic muscle discharges creating high noise levels and either a minimal or non-existent peak.

Beyond the individual findings for each condition, we have demonstrated that these two measures - particularly peak prominence - can be used to reliably identify a PSD peak, as opposed to relying on visual inspection. Given that it answers the question “Is there a peak?”, this measure could be a valuable tool for precisely defining a tremulous disorder as tremor.

### Frequency of peak activity

This measure identifies the predominant frequency at which muscle discharges occur. As expected from previous studies ^1, 10, 11^, there is no significant frequency difference between ET and DT, while OT shows a significantly higher frequency (Figure 3, panel C) ^1^. Interestingly, myoclonus results indicate not only a wide frequency range (8-40 Hz), but also the presence of very high frequencies. The most remarkable aspect is that these frequencies overlap with the beta frequency band, which is typically associated with high cortico-muscular coherence observed in cortical myoclonus ^6, 16, 17^.

### Peak width

The width of the peak activity defines the range of dominant frequencies. A wider peak width suggests that muscles can synchronously discharge across multiple frequencies rather than a single one, resulting in a broader, “bell-shaped” peak in the PSD rather than a sharp peak. This measure reflects the range of frequencies that contribute, with varying power, to tremor or myoclonus, offering insight into whether the phenomenon is driven by a single or multiple oscillatory activities, or a single oscillator influenced by interfering noise that alters the tremor frequency.

Our results indicate that myoclonus exhibits a larger peak width, likely due to irregular, multifrequency muscle discharges typical of arrhythmic phenomena, where no single dominant frequency emerges (Figure 3, panel D). A similar, broad peak width with more distinct peak prominence has been observed in cortical tremor, a form of rhythmic cortical myoclonus resembling tremor but sharing the same pathophysiology as cortical myoclonus ^5^. There has been debate over whether the cortical tremor spectrum is best described by a “frequency band” rather than a distinct “frequency peak,” given its typically small peak prominence (around 16 Hz) and broad peak width ^5, 18–20^, and our measures may help to clarify this.

DT also shows a broader peak width compared to ET and OT (Figure 3, panel D), suggesting higher frequency variability in DT. This characteristic has been proposed based on clinical observations of DT’s jerkiness and has been previously assessed using other measures, such as cycle-to-cycle frequency variability in the time domain or the tremor stability index, which captures frequency variability by examining the interquartile range of instantaneous frequency changes ^11, 21, 22^.

Comparable results to ours were achieved using similar measures of tremor powers, namely half-width power (HWP) and the peak at half-peak power (FWHM) ^10, 11^. The former quantifies tremor power by measuring the area under the curve between the rising and falling edges of the peak at half-peak power, while the latter provides a measure of frequency, specifically capturing the range of frequencies involved ^11^, which corresponds to our peak width. Similar to our results, FWHM was significantly higher in DT compared to ET, while HWP was lower ^11^. This finding suggests the presence of multiple oscillators with irregularities in both amplitude and frequency in DT or that superimposed dystonic contractions could disrupt the rhythmicity of the tremor.

In conclusion, the width of peak activity provides a measure of “regularity,” answering the question: “Is this tremor or myoclonus more or less regular?”. This measure could be particularly valuable, as studies suggest that more regular tremors may respond better to non-invasive central stimulation compared to irregular ones ^2, 3, 23^.

### Number of harmonics

The presence of harmonics has been observed in the PSD of tremor signals, particularly in studies on tremor in PD ^24–26^. The functional significance of these harmonics remains under debate. It is possible that they reflect true physiological phenomena, as previous studies have found significant cortico-muscular and cortico-cortical coherence at double the tremor frequency in PD and voluntarily simulated tremor ^25, 26^. This interpretation is further supported by the observation of different scalp topography of cortico-muscular coherence for the basic and first harmonic frequencies in PD tremor ^27^. While not directly tested here, this would support the presence of different neural generators in OT and ET, which could help differentiate them from DT and myoclonus, similar to what was reported previously between ET and PD tremor ^28^. Another possible explanation is that harmonics arise from sensory feedback from mechanoreceptors activated by muscle contractions associated with tremor ^25^. According to this view, the clearer and more numerous harmonics found in OT and ET in the present study may be a result of stronger and more synchronous tremor discharges, as demonstrated by the larger peak prominence and PB ratio found in these conditions, compared to DT and myoclonus.

The EMG signal itself may also contribute to harmonics in the PSD, potentially due to the non-linearity of tremor waveforms ^24, 25, 29^. It is important to note that the steepness of tremor discharges can have physiological meaning. The sum of individual motor unit action potentials correlates linearly with the neural drive to the muscle ^30^. The higher the level of motor unit synchronization, the steeper the rise and fall of the EMG bursts related to tremor ^30–32^. It has been suggested that the amplitude and number of harmonics in the EMG PSD depend on the steepness of the rising and falling edges in tremor discharges ^33^. In this view, our findings suggest that EMG discharges in OT are driven by the activity of a highly synchronous neural generator, while the descending activity in ET, DT, and myoclonus is more temporally dispersed.

### Conclusion

In this study, we demonstrated that our five outcome measures can objectively characterise tremulous involuntary movements, determining whether a movement qualifies as tremor, its regularity, and its predominant frequency or frequency range. Individual measures were able to differentiate OT, ET, DT, and myoclonus to different degrees. Combining all measures in a linear discriminant analysis allowed us to effectively separate these four movement disorders, providing significant segregation across them, with the exception of ET and DT (figure 5).

The inclusion of OT, ET, DT, and myoclonus was based on their distinct clinical features. We hypothesized that these disorders represent a continuum, ranging from a highly regular, single-frequency-driven phenomenon (OT) to an irregular, multifrequency phenomenon (myoclonus). Our results support this hypothesis, positioning OT and myoclonus at opposite ends of the spectrum, with ET and DT lying in between. The partial overlap in certain features, such as peak frequency and PB ratio, which lacked significant differences, may explain the limited accuracy of the discriminant function in separating ET and DT. By focusing exclusively on spectral features that demonstrated statistically significant differences between the two groups - specifically, peak prominence and peak width - DT and ET could be classified with high accuracy. The area under the ROC curve values were 0.905 for peak prominence and 0.963 for peak width, highlighting their strong discriminative power.

The presence of some overlap between DT and ET is not surprising. This might be due to diagnostic challenges ^34, 35^ or the broad inclusion of ET cases, including those classified as ET plus and questionable dystonia that may have clustered with DT cases. Nevertheless, we believe that these findings overall point to a potential pathophysiological link between ET and DT. While peak frequency and PB ratio are similar between the two, ET exhibits higher peak prominence and lower peak width compared to DT, suggesting that DT is characterized by greater background muscle activity and more variable frequencies, likely due to the influence of underlying dystonia. These differences may reflect slightly different physiological mechanisms: while both the cerebellum and basal ganglia are implicated in ET and DT ^11, 36–38^, a predominance of cerebellar activity may lead to the more regular, single-frequency tremor seen in ET, whereas a stronger influence of basal ganglia activity may result in more irregular, multifrequency tremor typical of DT. This interpretation aligns with previous research ^36, 39, 40^.

Our findings should be interpreted with caution due to the retrospective design of the study and the lack of in-depth phenotyping, such as differentiation of ET plus. Nonetheless, we believe our results provide valuable insights into tremor and cortical myoclonus characterisation, especially in differentiating ET from DT, potentially aiding clinical diagnosis and advancing understanding of the pathophysiology of these disorders.

## Data Availability

All data produced in the present study are available upon reasonable request to the authors

## Acknowledgements

None.

## Authors’ Roles

**Design:** AL, LR

**Execution:** AL, BH, CC, KH

**Analysis:** LR

**Writing:** AL, LR

**Editing of final version of the manuscript:** JCR, KPB

## Financial Disclosures of all authors (for the preceding 12 months)

KPB and AL are supported by EPSRC and MRC under the NEUROMOD+ Network (EP/W035057/1)

KPB receives grant support from Horizon 2020 EU grant 63482, honoraria/financial support to speak/attend meetings from GSK, Boehringer-Ingelheim, Ipsen, Merz, Sun Pharma, Allergan, Teva, Lundbeck and Orion pharmaceutical companies, royalties from Oxford University press.

**Supplementary Figure.** ROC curves to investigate discrimination accuracy between ET and DT according to peak prominence (panel A) and peak width (panel B). Red circles indicate optimal operating points (0.167, 0.875 for peak prominence; 0.075, 0.928 for peak width).

