## Supplementary figures and images for "Defining the rhythm: a new method to classify tremor and myoclonus"

### Supplementary figure

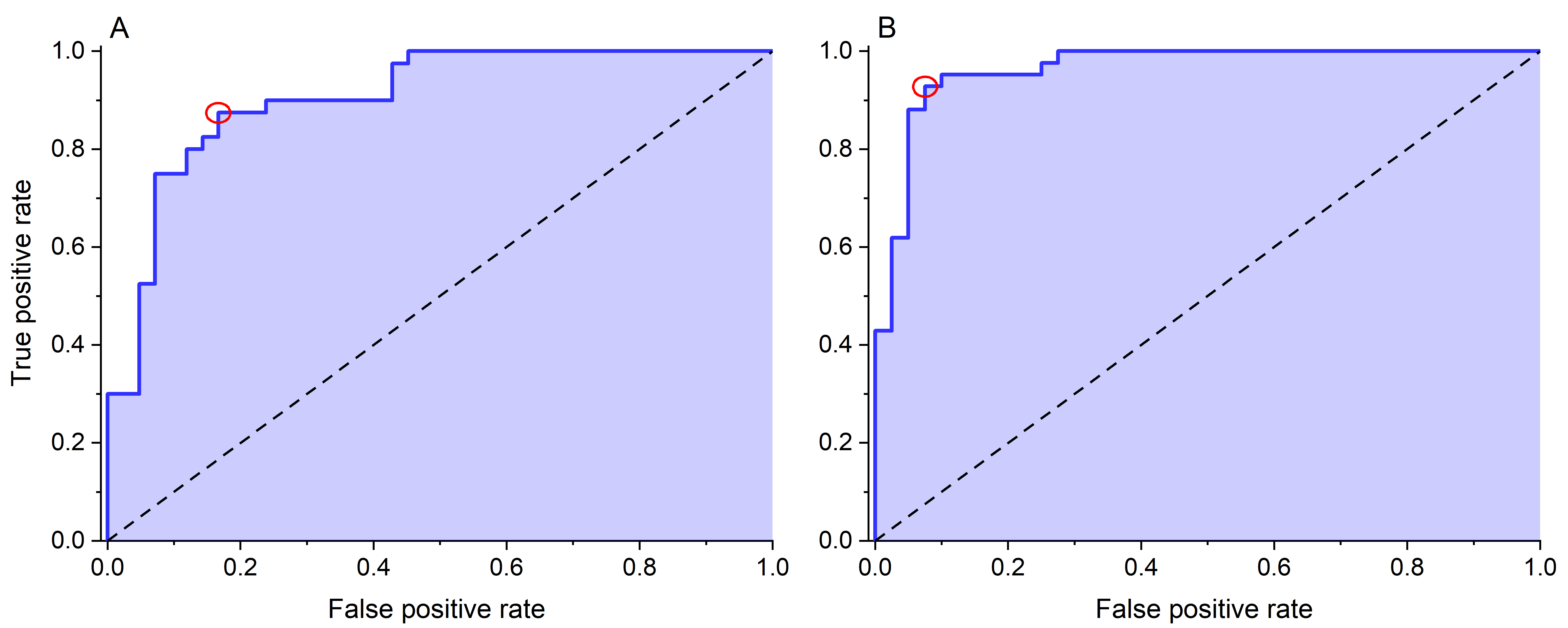
