## Supplementary materials for "Defining the rhythm: a new method to classify tremor and myoclonus"

EMG recording

EMG were recorded from a single channel by means of 10/20 mm diameter Ag/AgCl cup/pre-gelled adhesive electrodes and sampled at 5 kHz with a CED 1401 A/D laboratory interface (Cambridge Electronic Design, Cambridge, UK) or Micromed SD Flexi Plus laboratory interface (Micromed S.p.A., Mogliano Veneto, Italy). Electrodes were placed on the muscle most visibly affected by tremor or myoclonus, arranged in a belly-tendon montage. Muscle activity recordings were obtained under conditions that maximally activated it: standing for OT, holding arms outstretched for ET and DT, and during specific activation conditions associated with myoclonus. Each recording lasted approximately 90 seconds. Offline processing of signals included band-pass filtering between 1 and 1000 Hz and band-stop filtering between 48 and 52 Hz using a zero-phase, fourth-order Butterworth filter.
